# Superior long-term patency of no-touch vein graft compared to conventional vein grafts in over 1500 consecutive patients

**DOI:** 10.1101/2024.02.19.24303064

**Authors:** Gabriele Ferrari, Richard Loayza, Ava Azari, Håkan Geijer, Yang Cao, Roland Carlsson, Leif Bojö, Ninos Samano, Domingos Souza

**Affiliations:** University Health Care Research Centre, Faculty of Medicine and Health, Örebro University, Örebro, Sweden; Department of Cardiology and Cardiothoracic Surgery, Blekinge Hospital, Karlskrona, Sweden; Department of Radiology, Faculty of Medicine and Health, Örebro University, Örebro, Sweden; Clinical Epidemiology and Biostatistics, School of Medical Sciences, Faculty of Medicine and Health, Örebro University, Örebro, Sweden; Svensk PCI AB, Regional Hospital of Karlstad, Karlstad, Sweden; Clinical Physiology Division, Regional Hospital of Karlstad, Karlstad, Sweden

## Abstract

**Objectives:** To evaluate the long-term angiographic patency of saphenous vein grafts (SVG) harvested using the no-touch technique compared to the conventional technique.

**Methods:** This was a single-center, retrospective, cohort study. The inclusion criteria were individuals who underwent a CABG (coronary artery bypass grafting) between January 1995 and July 2020, and who successively needed a clinically-driven angiography. The primary endpoint was long-term patency. The secondary endpoints were differences in patency based on sub-group analysis (single vs. sequential graft, divided by target vessel).

**Results:** The study included 1520 individuals (618 no-touch, 825 conventional and 77 arterial grafts). The mean clinical follow-up time was 8.4 years ± 5.5 years. The patency per patient was 70.7% in the no-touch grafts vs. 46.7% in the conventional grafts (p < 0.001, OR = 2.8). The graft patency was 75.9% in the no-touch grafts vs. 62.8% in the conventional grafts (p < 0.001, OR= 1.8).

**Conclusions:** The no-touch vein grafts were associated with statistically significantly higher patency at long-term compared to the conventional grafts.

**Clinical Trial Registration:** NCT04656366, 7 December 2020

**Clinical Perspective:**

- What is new?

➣ The largest follow-up of the patency of no-touch vein grafts in the international literature.
➣ Patients with a no-touch vein graft had significantly better patency (p < 0.001) at mean follow-up of more than 8 years.
- What are the clinical implications?

➣ Consequent reduction in cardiovascular events after no-touch vein graft.
➣ Benefits at individual level due to fewer episodes of re-angina and myocardial infarction, and fewer coronary interventions.
➣ Benefits at community level due to fewer re-hospitalizations and a reduction in healthcare costs.

## Introduction

Ischemic heart disease is the leading cause of death worldwide, accounting for almost 8.9 million deaths in 2019 ^1^. The main treatment options are percutaneous coronary intervention (PCI), coronary artery bypass grafting (CABG) and, in some cases, both. The choice of treatment depends on the clinical presentation, grade of disease, type of lesion, and patient comorbidities. Guidelines ^2–4^ recommend CABG in cases of multivessel disease or left main stem, and/or diabetes mellitus, to improve survival and reduce the risk of major adverse cardiac events (MACE) ^5–9^, which are defined as death, myocardial infarction (MI), in-stent restenosis, and/or repeated target vessel revascularization (TVR). The grafts used, especially vein grafts, are subject to remodelling and progressive intimal hyperplasia and atherosclerosis, which may lead to graft stenosis or occlusion and thereby limit the graft patency ^7^. These events occur more frequently and rapidly in saphenous vein grafts (SVGs) compared to left internal thoracic artery (LITA) or other arterial grafts ^8, 9^. Graft patency after CABG is a major determinant of clinical prognosis and long-term survival ^7^.

The no-touch (NT) technique ^10^ differs from the conventional technique in that it is associated with less endothelium damage during the harvesting procedure ^11, 12^ and leads to reduced neo-intimal hyperplasia and subsequent atherosclerosis in the long term ^13, 14^.

The aim of this study was to evaluate the long-term angiographic patency of saphenous vein grafts (SVG) harvested using the NT technique compared to the conventional technique.

This is a single-center, retrospective, cohort study. The study complied with the STROBE (STrengthening the Reporting of OBservational studies in Epidemiology) statement and the STROBE checklist was used ^15^.

## Material and Methods

### Data collection

This single-center study included all individuals treated with a CABG between January 1, 1995 and June 30, 2020 at our Cardiothoracic Department and who successively needed a clinically driven-angiography. The study was a single-center study involving a center with long experience with the NT technique having used it for harvesting the saphenous vein since 1990. The study was performed in accordance with the Declaration of Helsinki. The Regional Ethical Review Board approved the study (Dnr 2020/04168).

The standard operation in our center has been to anastomise the LITA to the LAD (left anterior descending) and vein grafts to the other target vessels; in some cases, total arterial revascularization (A) occurred. The SVG was harvested either with the NT or with the C technique, as previously described ^16^. The type of technique used was chosen by the surgeon at the time of operation based on individual assessment and, therefore, without randomization. The angiographies were performed in two cardiology departments and were clinically driven, being due to either angina or myocardial infarction. No planned angiographic follow-up was performed. No exclusion criterion was defined.

The grade of stenosis and its significance was defined according to the 2021 American Heart Association for coronary artery revascularization ^17^. A limit of 70% was set for the non-left main stenosis and 50% for the left main stenosis or for graft stenosis; in case of doubt, the severity of the lesion was confirmed using the FFR (fractional flow reserve) or the iFR (instantaneous wave-free ratio).

The operative data (CABG) and the procedural data were collected with the help of an intervention-related quality register (the Swedish cardiological and cardiosurgical intervention register Swedeheart) ^18^. The operative data regarding the type of vein graft, the number of anastomoses, and the target vessel for the peripheral anastomosis were extracted and double checked by reviewing records in the clinical software of the two hospitals involved.

### Statistical methods

The primary endpoint was defined as long-term patency.

Secondary outcomes were the differences between single grafts and sequential grafts, and the different territories fed by the stenosed vein graft based on sub-group analyses.

Descriptive statistics were calculated as means ± standard deviations (SD) for normally distributed variables, as median with interquartile range (IQR) for non-normally distributed variables, and as count with percentage for categorical variables.

The Chi-squared test (or Fisher’s exact test for an expected count lower than 5) was used to compare categorical proportions between the two groups (NT and C). The unpaired t-test was performed to compare continuous variables and the Mann-Whitney U-test was used in cases of non-normally distributed data.

Statistical analyses were performed using SPSS version 27.0 (IBM, Armonk, NY, USA).

## Results

Between January 2006 and July 2020, 1571 individuals who had previously been treated with CABG needed an angiography (Figure 1). Fifty-one were lost at follow-up due to the inability to access the surgical reports (i.e., microfilm not available, journal lost, data protection).

**Figure 1:**
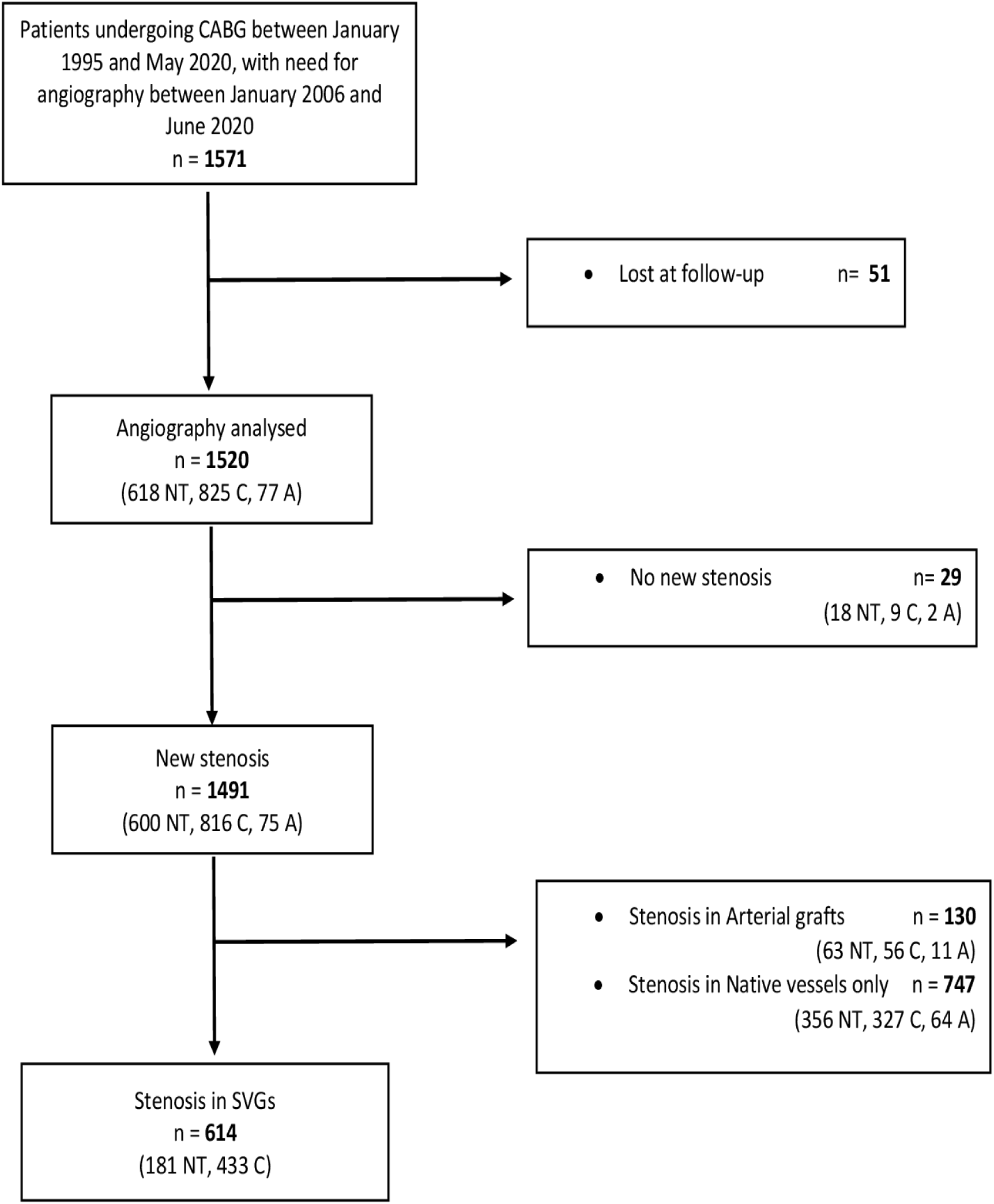
STROBE flowchart of the individuals included in the study. A: arterial graft alone; C: conventional graft; CABG: coronary artery bypass grafting; NT: no-touch graft; PCI: percutaneous coronary intervention; SVG: saphenous vein graft.

The study included 1520 consecutive patients (618 NT, 825 C, 77 A) who needed a postoperative angiography (see Figure 1 and Table 1) with a mean follow-up time of 8.4 ± 5.5 years and up to 24.7 years. The study analysed 2564 venous grafts (908 NT, 1656 C), Table 2.

**Table 1:** Graft characteristics of the patients in the total population.

| Patient characteristics | N (%) |
| --- | --- |
| No-touch | 618 (40.7) |
| Conventional | 825 (54.3) |
| LITA to LAD | 1273 (83.8) |
| RITA or Radial | 144 (9.5) |
| Total Arterial | 77 (5.1) |
| No-touch single grafts | 203 (13.4) |
| No-touch sequential grafts | 415 (27.3) |
| Conventional single grafts | 450 (29.6) |
| Conventional sequential grafts | 375 (24.7) |

**Table 2:** Vein graft characteristics of the two groups.

| **Patient characteristics** | **No-touch** | **Conventional** |
| --- | --- | --- |
| No. total grafts | 908 | 1656 |
| No. single grafts | 463 | 1247 |
| No. sequential grafts | 445 | 409 |

In 29 cases, the angiography was only diagnostic and no new stenosis was detected. In 614 cases, stenoses were present in at least one vein graft. One hundred and thirty individuals presented with a stenosis in an arterial graft, and in 747 cases the new stenosis affected only the native vessels.

### Demographic, operative, and procedural characteristics

A total of 1443 individuals received an SVG at surgery (618 NT, 825 C). The demographic, operative and angiographic characteristics of this group are summarized in Table 3. The demographic characteristics of the two groups were similar and patients had a comparable age at the time of CABG (63.2 ± 8.6 years NT, 63.1 ± 21.1 years C; p = 0.893; Table 3); most of the patients were male (78.5%). The risk factor distribution showed a significantly higher rate of hypertension (89% NT vs. 83.3% C) and hypercholesterolemia (91.7% NT vs. 84% C) in the NT group (p < 0.001). Fewer patients with an NT SVG received a post-operative DAPT (dual anti platelet therapy), 35.4% NT vs. 44% C (p = 0.001).

**Table 3:** Demographic characteristics of the individuals who received a saphenous vein graft.

| Patient characteristics | No-touch | Conventional | p-value |
| --- | --- | --- | --- |
| No. of patients, n (%) | 618 (40.7) | 825 (54.3) |  |
| Mean age at CABG, years ± SD | 63.2 ± 8.6 | 63.1 ± 21.1 | 0.893 |
| Male, n (%) | 506 (81.9) | 626 (75.9) | 0.006 |
| Hypertension, n (%) | 550 (89.0) | 687 (83.3) | <0.001 |
| Hypercholesterolemia, n (%) | 567 (91.7) | 693 (84.0) | <0.001 |
| Diabetes mellitus, n (%) | 216 (39.8) | 284 (34.4) | 0.895 |
| Smoking, n (%) | 349 (56.5) | 440 (53.3) | 0.359 |
| Double platelet inhibitor therapy, n (%) | 219 (35.4) | 363 (44.0) | 0.001 |
| Creatinine median (Q1, Q3) | 85 (72, 102) | 88 (75, 105) | 0.004* |
| Dead at follow-up, n (%) | 170 (27.5) | 420 (50.9) | <0.001 |

\* Mann-Whitney U test was used. T-test was used for continuous variables and Chi-squared test for categorical variables. Values are presented as mean ± standard deviation for normal distributed variables, median (Q1, Q3) for non-normal distributed variables, or n (%). Ages and times are given in years. Creatinine is given in micromole/L.

The operative characteristics regarding number of peripheral anastomoses were similar in the two groups (Table 4). No differences regarding the indication for the new angiography were presented (p = 0.916), Table 4.

**Table 4:**
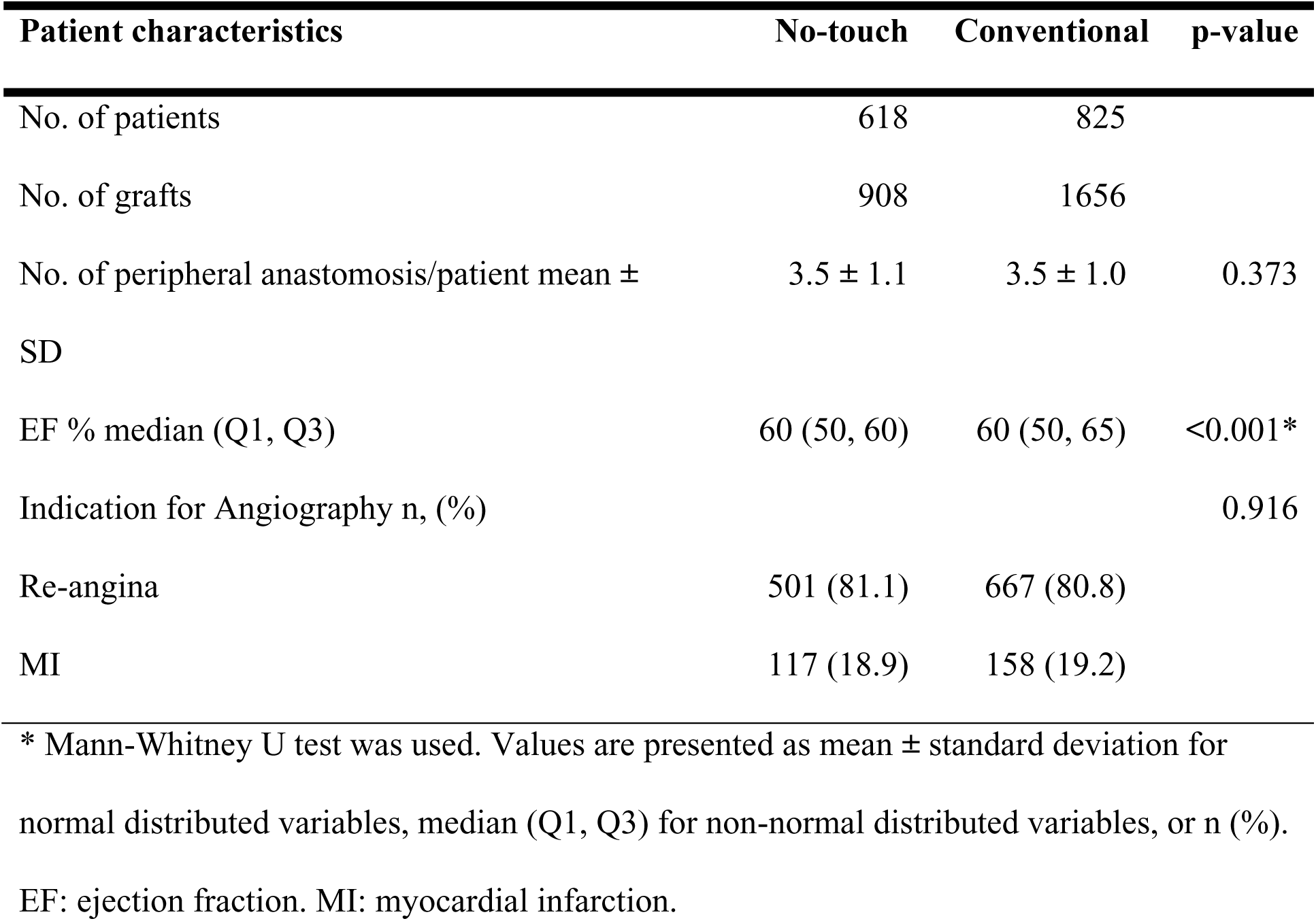
Operative and procedural characteristics of the individuals who received a saphenous vein graft.

| Patient characteristics | No-touch | Conventional | p-value |
| --- | --- | --- | --- |
| No. of patients | 618 | 825 |  |
| No. of grafts | 908 | 1656 |  |
| No. of peripheral anastomosis/patient mean ± SD | 3.5 ± 1.1 | 3.5 ± 1.0 | 0.373 |
| EF % median (Q1, Q3) | 60 (50, 60) | 60 (50, 65) | <0.001* |
| Indication for Angiography n, (%) |  |  | 0.916 |
| Re-angina | 501 (81.1) | 667 (80.8) |  |
| MI | 117 (18.9) | 158 (19.2) |  |

\* Mann-Whitney U test was used. Values are presented as mean ± standard deviation for normal distributed variables, median (Q1, Q3) for non-normal distributed variables, or n (%). EF: ejection fraction. MI: myocardial infarction.

The mean follow-up time was 5.5 ± 4.4 years for the individuals with an NT SVG and 10.7 ± 5.1 years for the individuals with a C SVG.

### Clinical follow-up

#### Primary endpoint

The total patency rate per patient was 70.7% (437/618) in the NT group and 46.7% (385/825) in the C group (p < 0.001), with an OR (odds ratio) of 2.8, Table 5.

**Table 5:** Patency rate per patient of no-touch and conventional vein grafts, and patency of arterial grafts divided by type of vein graft.

| Patient characteristics | No-touch | Conventional | p-value |
| --- | --- | --- | --- |
| No. of patients | 618 | 825 |  |
| Patency/patient of Vein Grafts, % | 70.7 | 46.7 | <0.001 |
| Patency/patient of Arterial grafts, % | 86.4 | 86.3 | 0.954 |
| Patency/patient of LITA, % | 98.4 | 95.9 | 0.006 |

Values are presented as %. LITA: left internal thoracic artery.

The patency rate per graft was 75.9% (609/908) in the NT group and 63.5% (1051/1656) in the C group (p < 0.001), with an OR of 1.8, Table 6.

**Table 6:** Patency in no-touch and conventional vein grafts divided by type of graft.

| **Patient characteristics** | **No-touch** | **Conventional** | **p-value** |
| --- | --- | --- | --- |
| No. of Grafts | 908 | 1656 |  |
| Graft Patency | 75.9 | 63.5 | <0.001 |
| Single | 75.6 | 62.8 | <0.001 |
| Sequential | 76.0 | 65.3 | <0.001 |

Values are presented as %.

#### Secondary endpoint

##### Single grafts vs. sequential grafts

In the analysis of the different types of graft received (single or sequential), there was a statistically significant higher patency in the NT group both for single grafts (75.6% NT vs. 62.8% C, p < 0.001) and for sequential grafts (76.0% NT vs. 65.3% C, p < 0.001), Table 6.

##### Territories fed by the stenosed vein graft

The subgroup analysis, divided by distal anastomosis, revealed statistically significantly better results for the NT grafts for each of the different distribution territories, Table 7. The patency of the NT SVGs was over 80% if the target vessel was a coronary of the left system, and around 70% if the target vessel was the right coronary artery (RCA). In contrast, the patency of the C SVGs was as low as 57% if the target vessel was the RCA, and never reached 80% in the left system. The differences between the NT and the C in the four different territories of distribution were significant even when analysing the single and the sequential grafts separately; however, the exceptions were the single grafts to the marginal artery (MA), the sequential ones to RCA, and both types to the left anterior descending (LAD) due to the limited number of included individuals, Table 7.

**Table 7:** Patency of no-touch and conventional vein grafts divided by territory of distribution and sub-divided by graft type (single or sequential)

| Patient characteristics | No-touch | Conventional | p-value |
| --- | --- | --- | --- |
| To LAD | 58/70 (82.9) | 45/66 (68.2) | 0.046 |
| Single | 30/36 (83.3) | 35/49 (71.4) | 0.201 |
| Sequential | 28/34 (82.4) | 10/17 (58.8) | 0.093* |
| To DA | 317/360 (88.1) | 353/519 (68.0) | <0.001 |
| Single | 44/54 (81.5) | 160/273 (58.6) | 0.002 |
| Sequential | 273/306 (89.2) | 193/246 (78.5) | <0.001 |
| To MA | 402/468 (85.9) | 529/673 (78.6) | 0.002 |
| Single | 90/104 (86.5) | 270/339 (79.6) | 0.115 |
| Sequential | 312/364 (85.7) | 259/334 (77.5) | 0.005 |
| To RCA | 285/404 (70.5) | 355/623 (57.0) | <0.001 |
| Single | 183/256 (71.5) | 307/549 (55.9) | <0.001 |
| Sequential | 102/148 (68.9) | 48/74 (64.9) | 0.543 |

Values are presented as n, (%). LAD: left anterior descending, DA: diagonal artery,
MA: marginal artery, RCA: right coronary artery. \* Fisher's exact test.

##### Patency of arterial grafts

The long-term patency of the total arterial grafts was 86.3% and of the LITA grafts 96.2%. The patency rates for arterial grafts were similar regardless of whether the patients received an SVG or not, Table 5.

## Discussion

This study presents the results from the largest long-term follow-up investigation of the qpatency of NT SVGs.

The main finding of this study was higher graft patency at follow-up of the NT grafts (75.9%), compared to the C grafts (63.5%); p < 0.001. The superiority of the NT SVG was shown even when the different types of grafts (single vs. sequential) and the different types of territory of distribution were analysed. It has to be mentioned that NT grafts have shorter follow-up compared to the C grafts; however, this is due to higher rates of restenosis in the native vessels and not in the SVGs themselves.

It is known that C SVGs have high risks of thrombosis, intimal hyperplasia, and early atherosclerosis, which all lead to graft occlusion rates of up to 50-60% after 10 years, according to most clinical records ^19–21^.

Our study showed similar results for the C grafts with a patency per patient of only 46.7% at a mean follow-up of 8.4 years. The NT grafts, on the other hand, present a patency per patient of more than 70% at follow-up (OR of 2.8) and of almost 76% per graft.

Several studies ^11, 13, 14, 16, 22–25^ have shown the superiority of NT SVGs compared to C SVGs on follow-up at a mean time up to 16 years ^14^. Previous studies have, however, had either a limited follow-up time of maximum 1 year ^22, 23, 25^ or included a limited number of patients (fewer than 300) ^11, 13, 14, 16, 23, 24^. The present study is the first to include both a large cohort, more than 1500 individuals, and a mean follow-up time of more than 5 years and up to almost 25 years. The higher long-term patency of NT SVGs may play an important role in reduced long-term MACE and re-hospitalization rates.

In our previous study ^26^ we noted a very limited number of patients with NT SVGs being treated with PCI compared to C (63 NT vs. 246 C). We also noted that almost 70% of patients who needed a coronary angiography after the CABG operation did not receive a PCI on their vein graft. The present study gives an explanation for these observations; more than half (57.6%) of the patients with an NT SVG who needed a postoperative angiography did not have a SVG stenosis, but presented with a new stenosis in the native vessels. This occurred in only 39.6% of patients with a C SVG.

Another interesting data is the higher percentage of sequential grafts in the NT group (49.3% vs. 24.7%). This is possible thanks to the qualities of the NT grafts that are much less prone to kinking when performing multiple anastomoses ^12^, comparing the C technique.

As previously mentioned, NT grafts have a shorter time to follow-up compared to C grafts, with only 23 cases (vs. 124) needing an angiography more than 15 years after the operation. This shorter follow-up time for NT grafts is due to a lower patency rate of native vessels in these individuals. A possible explanation is the statistically significant difference (35.4% vs 44.0%, p = 0.001) in the use of DAPT, with NT grafts receiving postoperative DAPT in fewer cases compared to C grafts. The benefits of DAPT after an acute coronary syndrome, in terms of reduced risk of restenosis and higher short- and long-term patency rates, are well known and supported by the international guidelines ^27^. Another possible explanation is that in the 1990’s, the C graft was used in most cases so these patients automatically have a longer follow-up time than the NT grafts.

The main limitation of this study is that it is non-randomized and the choice of graft type was dependent on the surgeon’s choice. Another limitation is the different follow-up times in the two groups.

## Conclusions

The results regarding the NT SVGs are encouraging, with statistically significantly higher patency at long-term follow-up compared to a C SVG, and with an almost 50% reduction in cases of vein graft stenosis long-term.

The results suggest an important reduction of cardiovascular events if the patients receive comparable medical treatment post-surgery and if the management of the risk factors follows the guidelines.

Randomized studies are needed to evaluate the performance and benefits of NT SVGs at both individual (better patency and fewer coronary interventions) and community (fewer re-hospitalizations) levels.

## Data Availability

https://classic.clinicaltrials.gov/ct2/show/NCT04656366

## Non-standard abbreviations and non-standard acronyms

A: arterial graft
BMS: bare metal stent
C: conventional
CABG: coronary artery bypass grafting
DA: diagonal artery
DAPT: double antiplatelet therapy
DES: drug-eluting stent
EF: ejection fraction
FFR: fractional flow reserve
iFR: instantaneous wave-free ratio
IQR: interquartile range
LAD: left anterior descending artery
LITA: left internal thoracic artery
MA: marginal artery
MACE: major adverse cardiac events
MI: myocardial infarction
NT: no-touch
OR: odds ratio
PCI: percutaneous coronary intervention
RCA: right coronary artery
RITA: right internal thoracic artery
SD: standard deviation
SVG: saphenous vein graft
TIMI: thrombolysis in myocardial infarction
TVR: target vessel revascularization

## Acknowledgements

We thank research assistant Ulf Graf for help with the database settings.

## Sourcing of Fundings

This work was supported by Region Örebro County through the regional research board (grant OLL-OLL-935188, 18 January 2020).

## Supplementary Files

Supplementary Table S1

Supplementary Table S2

Supplementary Table S3

## Author contribution statement

Gabriele Ferrari, Håkan Geijer, Ninos Samano, Domingos Souza, Roland Carlsson and Leif Bojö devised the original concept of this study. Gabriele Ferrari, Håkan Geijer, Ninos Samano, Domingos Souza, Roland Carlsson and Leif Bojö designed the study. Gabriele Ferrari, Richard Loayza and Ava Azari collected the patient data. Gabriele Ferrari, Richard Loayza and Ava Azari performed the statistical analysis. Yang Cao provided support for statistical analysis and result explanation. Gabriele Ferrari, and Yang Cao wrote the first draft of the manuscript. All authors reviewed the manuscript, contributed significantly to its critical review, and approved the final version.

